# Prevalence of diabetic foot ulcer and its association with duration of illness and residence in Ethiopia: a systematic review and meta-analysis

**DOI:** 10.1101/19003061

**Authors:** Henok Mulugeta, Fasil Wagnew, Haymanot Zeleke, Bekele Tesfaye, Haile Amha, Cheru Tesema Leshargie, Henok Biresaw, Getenet Dessie, Yihalem Abebe Belay, Tesfa Dejenie Habtewold

**Affiliations:** Lecturer of Nursing, Department of Nursing, College of Health Science, Debre Markos University Address: P.O. Box 269, Debre Markos, Ethiopia; Lecturer of Environmental Health, Department of Environmental Health, College of Health Science, Debre Markos University, Address: P.O. Box 269, Debre Markos, Ethiopia; Lecturer of Nursing, Department of Nursing, School of health science, College of Medicine and Health Science, Bahir Dar University, Address: P.O. Box 79, Bahir Dar, Ethiopia; Lecturer of Public Health, Department of Public Health, Debre Markos University Address: P.O. Box 269, Debre Markos, Ethiopia; Department of Epidemiology, University of Groningen, University Medical Center Groningen, Groningen, the Netherlands

**Keywords:** Diabetes mellitus, foot ulcers, prevalence, Ethiopia

## Abstract

**Background:** Diabetic foot ulcer (DFU), devastating complications of diabetes mellitus, is a major public health problem, and one of the leading reasons for hospital admission, amputations, and even death among diabetic patients in Ethiopia. Despite its catastrophic health consequences, the national burden of diabetic foot ulcer remains unknown in Ethiopia. Hence, the objective of this systematic review and meta-analysis was to estimate the national prevalence of diabetic foot ulcer and investigate the association with duration of illness and patient residence among diabetic patients.

**Methods:** We searched PubMed, Google Scholar, Cochrane Library, CINAHL, EMBASE, and PsycINFO databases for studies of diabetic foot ulcers prevalence that published from conception up to June 30, 2019. Quality of each article was assessed using a modified version of the Newcastle-Ottawa Scale for cross-sectional studies. All statistical analyses were done using STATA version 14 software for Windows, and meta-analysis was carried out using a random-effects method. The pooled national prevalence of diabetic foot ulcers was presented using a forest plot.

**Results:** A total of 10 studies with 3,029 diabetic patients were included. The pooled national prevalence of diabetic foot ulcers among Ethiopian diabetic patients was 11.27% (95% CI 7.22, 15.31%, I^2^=94.6). Duration of illness (OR: 3.91, 95%CI 2.03, 7.52, I^2^=63.4%) and patients’ residence (OR: 3.40, 95%CI 2.09, 5.54, I^2^=0.0%) were significantly associated with a diabetic foot ulcer.

**Conclusion:** In Ethiopia, at least one out of ten diabetic patients had diabetic foot ulcers. Healthcare policymakers (FMoH) need to improve the standard of diabetic care and should design effective preventive strategies to improve health care delivery for people with diabetes and reduce the risk of foot ulceration.

## Background

Diabetes is a chronic disorder characterized by hyperglycemia that occurs either due to insulin deficiency or resistance[1, 2] that has been increasing rapidly worldwide. According to the American Diabetes Association 2014 report, the global prevalence of diabetes among adults is 8.5%[2, 3]. In Africa, an estimated 12.1 million people were living with diabetes in 2010, and this figure is more likely to be 23.9 million by 2030[4].

Diabetes has several acute and chronic complications including diabetic foot ulceration. A diabetic foot, which is the formation of foot ulceration that manifested by neuropathy, ischemia, and infection, is one of the major public health problems affecting diabetic patients throughout the world[5, 6]. About 10% to 15% of diabetes patients are at risk of developing a foot ulcer during their lifetime[7]. The annual population-based incidence of diabetic foot ulceration ranges from 1.0% to 4.1%, and its prevalence among diabetic patients ranges from 4% to 10%[5, 8].

Diabetes foot problems are responsible for high morbidity and mortality of diabetes in Africa. In sub-Saharan Africa, the burden of a diabetic foot ulcer is increasing due to late diagnosis, poor awareness among patients, and poor access to diabetic care[9, 10]. The presence of foot ulcers in diabetes patient increases the risk of lower extremity amputations, worsens the physical, psychological, and social quality of life[11-13]. Furthermore, patients living with diabetic foot ulcers(DFUs) has a great economic burden and poor health-related quality of life[14].

Diabetic foot ulceration has been associated with various risk factors, such as longer duration of diabetes mellitus (DM), residence, lack of awareness, poor foot care practice, poor glycemic control, poor hygienic condition, foot trauma, peripheral neuropathy and presence of vascular diseases[15-17] [8] [18] [19]. Interestingly, most of the risk factors can be prevented easily with health education, careful foot care, and glycemic control[20].

Despite the huge burden of diabetic foot ulceration and it is one of the challenging health concerns in Ethiopia, the overall prevalence of diabetic foot ulcer among diabetic patients at the national level remains unknown. Therefore, the objective of this systematic review and meta-analysis was to estimate the national prevalence of foot ulcers among diabetic patients and investigate the association with duration of illness and residence in Ethiopia. The findings of this study will be important to provide valuable information regarding the burden of the diabetic foot and to inform healthcare policymakers for areas of improvement in diabetic care in Ethiopia.

## Methods

### Search strategy and selection criteria

This systematic review and meta-analysis were conducted following the Preferred Reporting Items for Systematic Reviews and Meta-Analyses (PRISMA) guideline[21]. We searched articles reporting the prevalence of diabetic foot ulcers among diabetic patients from PubMed, Cochrane Library, Google Scholar, CINAHL, and EMBASE databases using the following search terms: “prevalence”, “diabetic”, “diabetic foot”, “ulcers”, “diabetic foot ulcers”, “patients”. Boolean operators like “AND” and “OR” were used to combine search terms. EndNote (version X8) software for Windows was used to search, download, organize, and cite the related articles. The reference lists of eligible articles were also searched to retrieve additional relevant articles.

All published and unpublished studies that report the prevalence of diabetic foot ulcers among diabetic patients in Ethiopia were included. Studies, which investigated the association between duration of illness and patient residence with foot ulceration, were also included. Only these two factors such as diabetic duration and patient residence have been similarly assessed across the included studies. Therefore, we estimate the pooled effects of these factors to determine their association with diabetic foot ulcers. The search for published studies was restricted to articles published in English language. Furthermore, due to limited literature, we reviewed all publications irrespective of publication year. Nevertheless, related articles which failed to report the prevalence of diabetic foot ulcers were excluded. The articles were searched from March 2019 to June 2019.

### Data extraction

All the downloaded studies were independently screened for inclusion by two authors (HM and GD). From each included study, information on the name of the first author, year of publication, study area (region), health facility, study design, sample size, response rate, prevalence of diabetic foot ulcers and determinant factors (Diabetic duration and patient residence) were extracted using a pre-piloted template prepared in a Microsoft Excel spreadsheet. H.M. and H.A. conducted the primary data extraction and then B.T., F.W., and H.Z. examined the extracted data independently. Any disagreement and inconsistencies were resolved by discussion.

### Quality assessment

We used the modified version of the Newcastle-Ottawa Scale for cross-sectional studies adapted from Modesti et al to assess the methodological and other quality of each relevant article. The key criteria in the Newcastle–Ottawa scale are representativeness of the sample, response rate, measurement tool used, comparability of the subject, appropriateness of the statistical test used to analyze the data[22]. Two authors (B.T, and H.B) independently assessed the quality of each article. Any disagreement was resolved through discussion and consensus.

### Statistical Analysis

A random-effects model using the DerSimonian and Laird method[23] was employed to estimate the pooled national prevalence and OR of risk factors with a 95% confidence interval (CI). The result was presented using a forest plot. Heterogeneity across the studies was assessed by Cochrane’s Q test and I^2^ statistic. I^2^ statistic ranges from 0 to 100 percent. We used the accepted I^2^statistic > 50% to define significant heterogeneity across the included studies[24, 25]. Moreover, we explored the source of between-study heterogeneity using meta-regression analysis. A funnel plot was used for visual assessment of publication bias. Asymmetry of the funnel plot indicates the presence of publication bias[26, 27]. Egger’s tests were also used to assess the significance of publication bias with a p-value less than 0.05[28]. Finally, a sensitivity analysis was performed to assess whether the pooled prevalence estimates were influenced by individual studies. All data manipulation and statistical analyses were performed using Stata version 14.0 software for Windows.

## Results

### Study selection

The online search process initially yielded 1079 articles. Of which 21 articles duplicate records were identified and removed. After reviewing the title and abstract, we excluded 1016 irrelevant articles. From the remaining 42 articles, 32 articles were excluded since they failed to meet eligibility criteria and having poor quality based on the Newcastle-Ottawa Scale. Finally, a total of 10 relevant articles with 3,029 diabetic patients were included in the meta-analysis (Figure 1).

**Figure 1:**
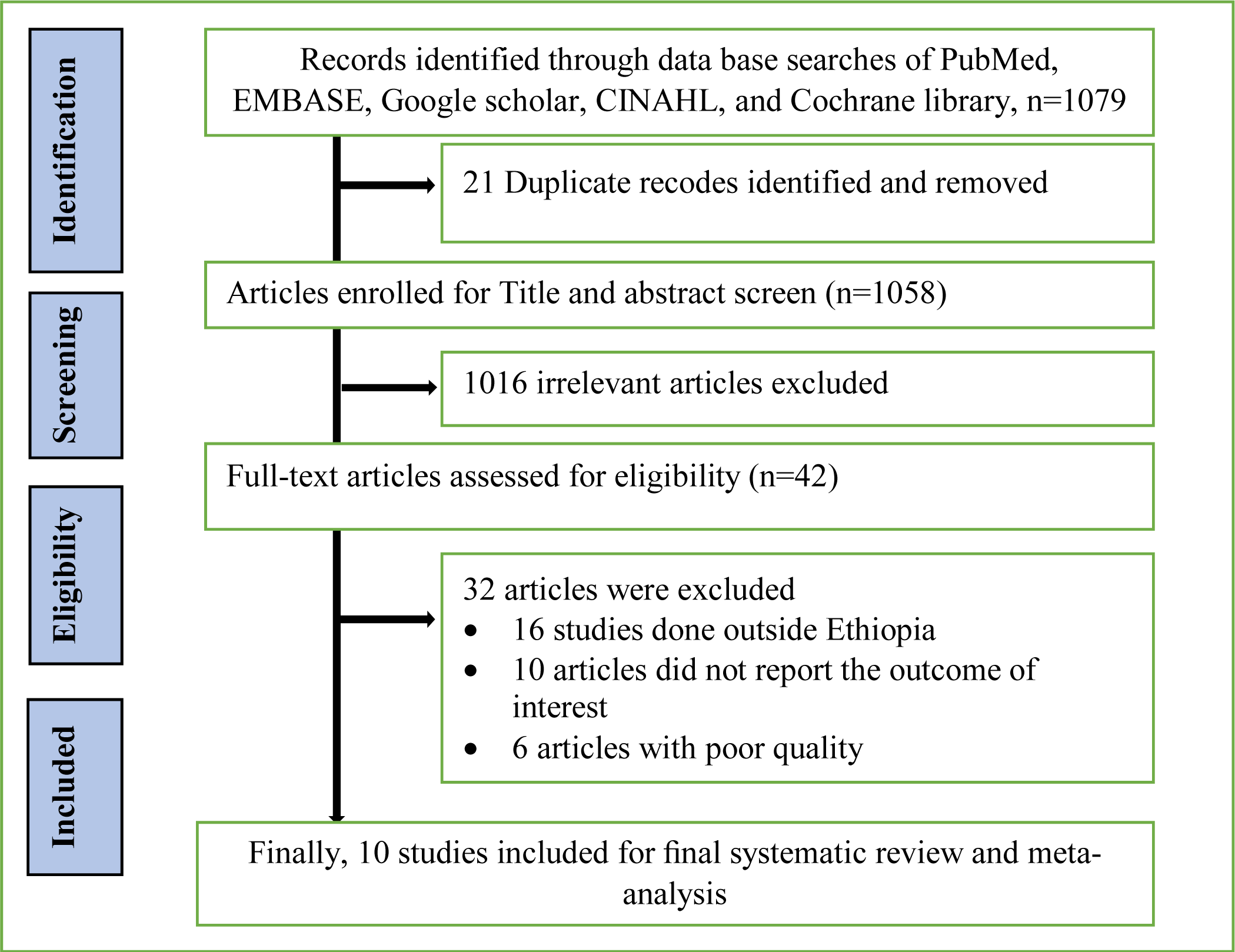
PRISMA Flowchart diagram of the study selection.

### Study characteristics

The studies were conducted from 2000 to 2017 in different regions of the country. Of the 10 studies, three were conducted in Addis Ababa[29-31], three were in Amhara[32-34], two were in Oromia[35, 36], and the other two[37, 38] were from other regions. All the included studies were cross-sectional by study design and their sample size ranged from 198 to 724 patients (Table1).

**Table 1:** Characteristics of studies included in the meta-analysis of foot ulcers among diabetic patients in Ethiopia

| No | Author/s | Publication Year | Study area, Region | Health Facility Name | Study Design | Sample Size | Prevalence % (95%CI) | Risk factors | Funding |
| --- | --- | --- | --- | --- | --- | --- | --- | --- | --- |
| 1 | Yimam A.[31] | 2017 | Addis Ababa | Black Lion Hospital | Cross-sectional | 198 | 26.0 (19.9,32.1) | -Residence<br>-Duration of diabetes | Addis Ababa University |
| 2 | Ejigu A.[30] | 2000 | Addis Ababa | Menelik Hospital | Cross-sectional | 283 | 1.70 (0.20,3.20) | ----- | ----- |
| 3 | Adem A. et al[29] | 2011 | Addis Ababa | Black Lion and St. Paul's Hospitals | Cross-sectional | 724 | 9.7 (7.54,11.8) | ----- | Addis Ababa University |
| 4 | Lebeta KR. et al[33] | 2017 | Bahir Dar, Amhara | Felege Hiwot Referral Hospital | Cross-sectional | 344 | 21.2 (16.9,25.5) | -Duration of diabetes | Bahir Dar University |
| 5 | Abejew AA. et al[32] | 2015 | Dessie, Amhara | Dessie Referral Hospital | Cross-sectional | 216 | 4.4 (1.7,7.1) | ----- | Wollo University |
| 6 | Mariam TG. et al[34] | 2017 | Gondar, Amhara | University of Gondar Hospital | Cross-sectional | 279 | 13.6 (9.6,17.6) | -Residence | University of Gondar |
| 7 | Worku D. et al[36] | 2010 | Jimma, Oromia | Jimma University Specialized Hospital | Cross-sectional | 305 | 4.6 (2.3,6.9) | ----- | Jimma University |
| 8 | Tilahun A. et al[35] | 2017 | Jimma, Oromia | Jimma University Specialized Hospital | Cross-sectional | 236 | 8.5 (4.9,12.1) | ----- | Jimma University |
| 9 | Gebrekiros K. et al[38] | 2015 | Mekelle, Tigray | Ayder Specialized Hospital | Cross-sectional | 228 | 12.0 (7.8,16.2) | ----- | Mekelle University |
| 10 | Deribe B. et al[37] | 2014 | Arba Minch, SNNP | Arba Minch General Hospital | Cross-sectional | 216 | 14.8 (10.1,19.5) | -Residence<br>-Duration of diabetes | Jimma University |

### Prevalence of diabetic foot ulcers

The pooled national prevalence of diabetic foot ulcers among diabetic patients was 11.27% (95% CI (7.22, 15.31%), I^2^=94.6). The overall pooled effect size of foot ulceration in diabetes mellitus patients presented using forest plot (Figure2).

**Figure 2:**
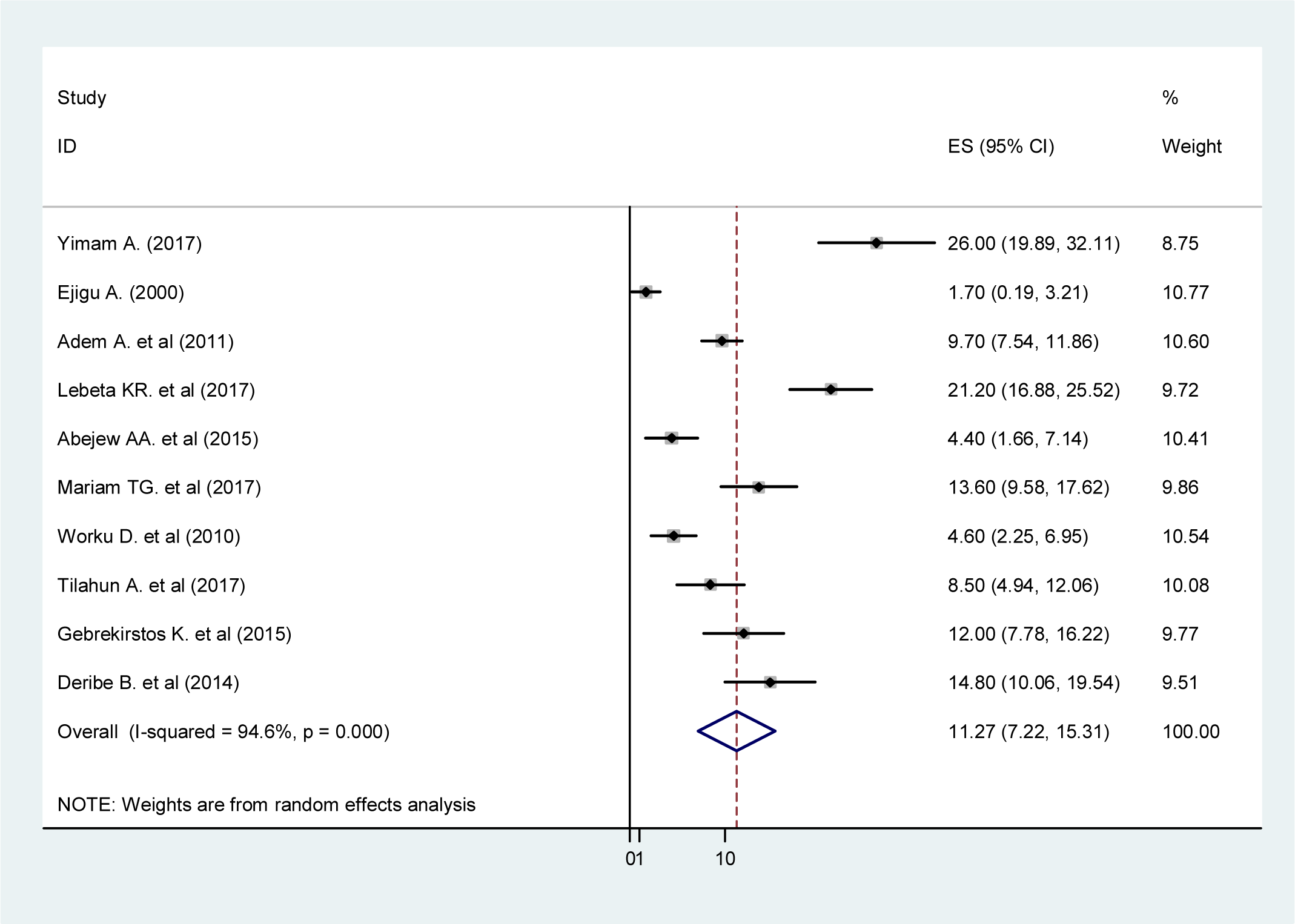
Forest plot showing the pooled prevalence of diabetic foot ulcers.

### Assessment of Heterogeneity

In this meta-analysis, the presence of heterogeneity was checked by I^2^ statistics. The result of this meta-analysis using the random-effects model revealed a high heterogeneity across the included studies (I^2^ = 94.6%, P=0.001). The sources of variation were assessed using a meta-regression model using publication year and sample size as covariates. The result of the meta-regression analysis showed that both covariates were not statistically significant for the presence of heterogeneity (Table 2).

**Table 2:**
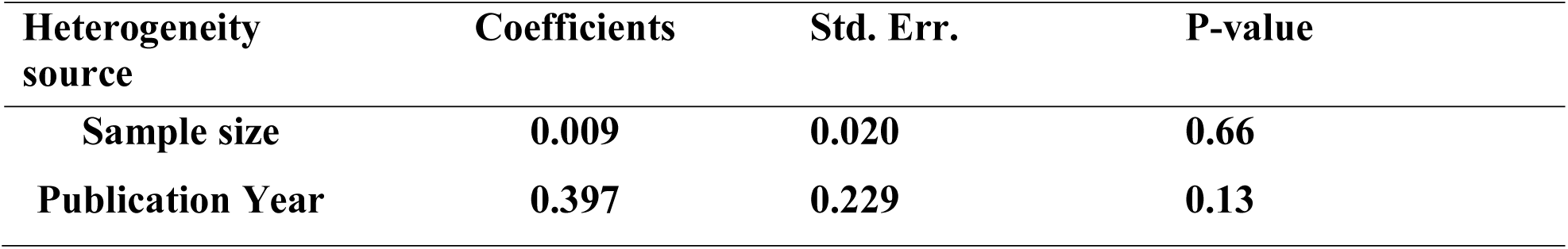
Meta-regression analysis of factors with heterogeneity

| Heterogeneity source | Coefficients | Std. Err. | P-value |
| --- | --- | --- | --- |
| Sample size | 0.009 | 0.020 | 0.66 |
| Publication Year | 0.397 | 0.229 | 0.13 |

### Assessment of publication bias

Publication bias was examined by using a funnel plot and Egger’s test. Visual inspection of the funnel plot suggests asymmetry, as three studies lay on the left side and seven studies on the right side of the line (Figure 3). Besides, asymmetry of the funnel plot was statistically significant as evidenced by Egger’s test (P=0.001), which indicated the presence of publication bias. Furthermore, trim and fill analysis, which is a nonparametric method for estimating the number of missing studies[39], was performed to reduce and adjust publication bias(Figure 4).

**Figure 3:**
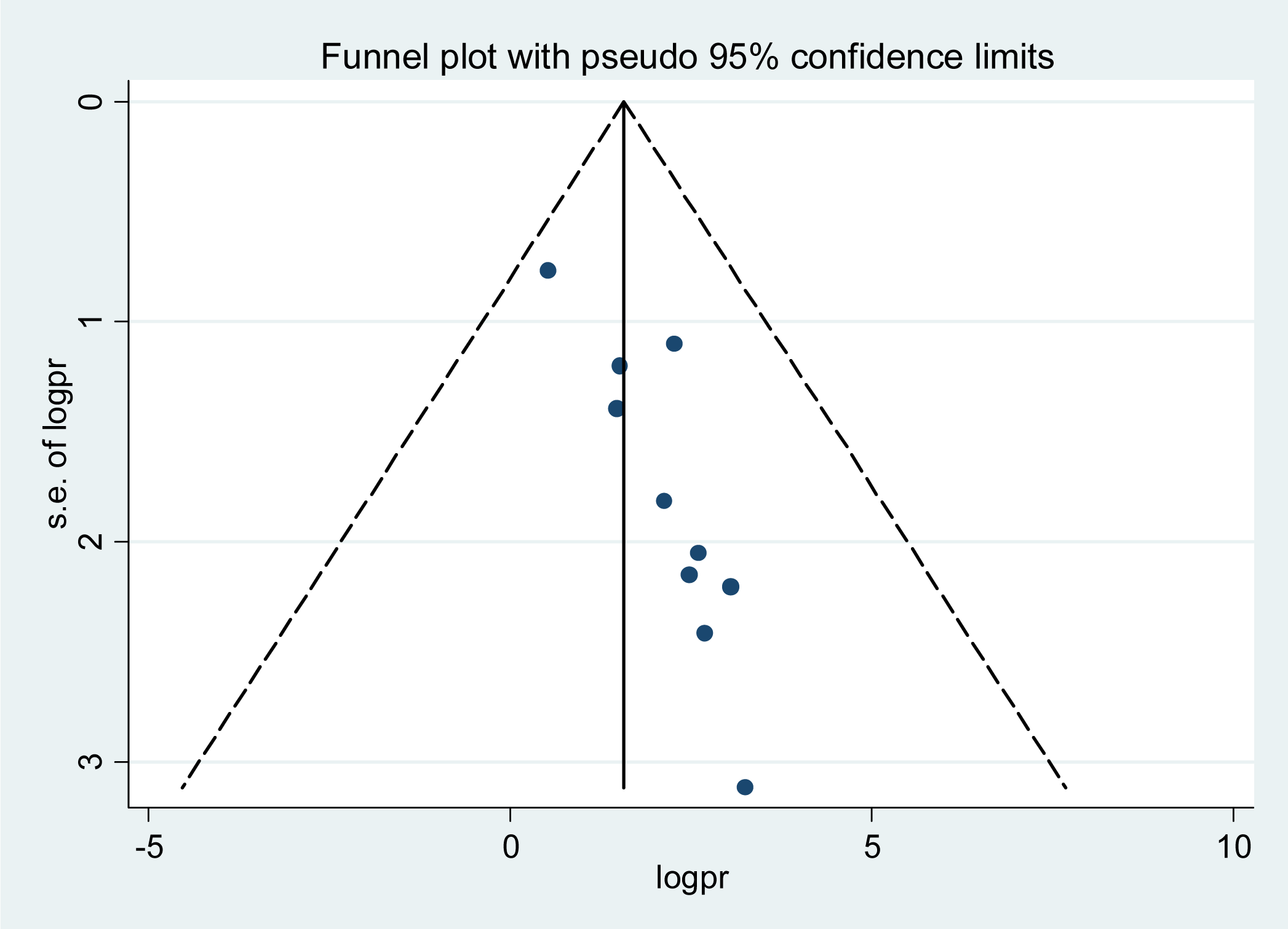
Funnel plot to test the publication bias of the 10 studies.

**Figure 4:**
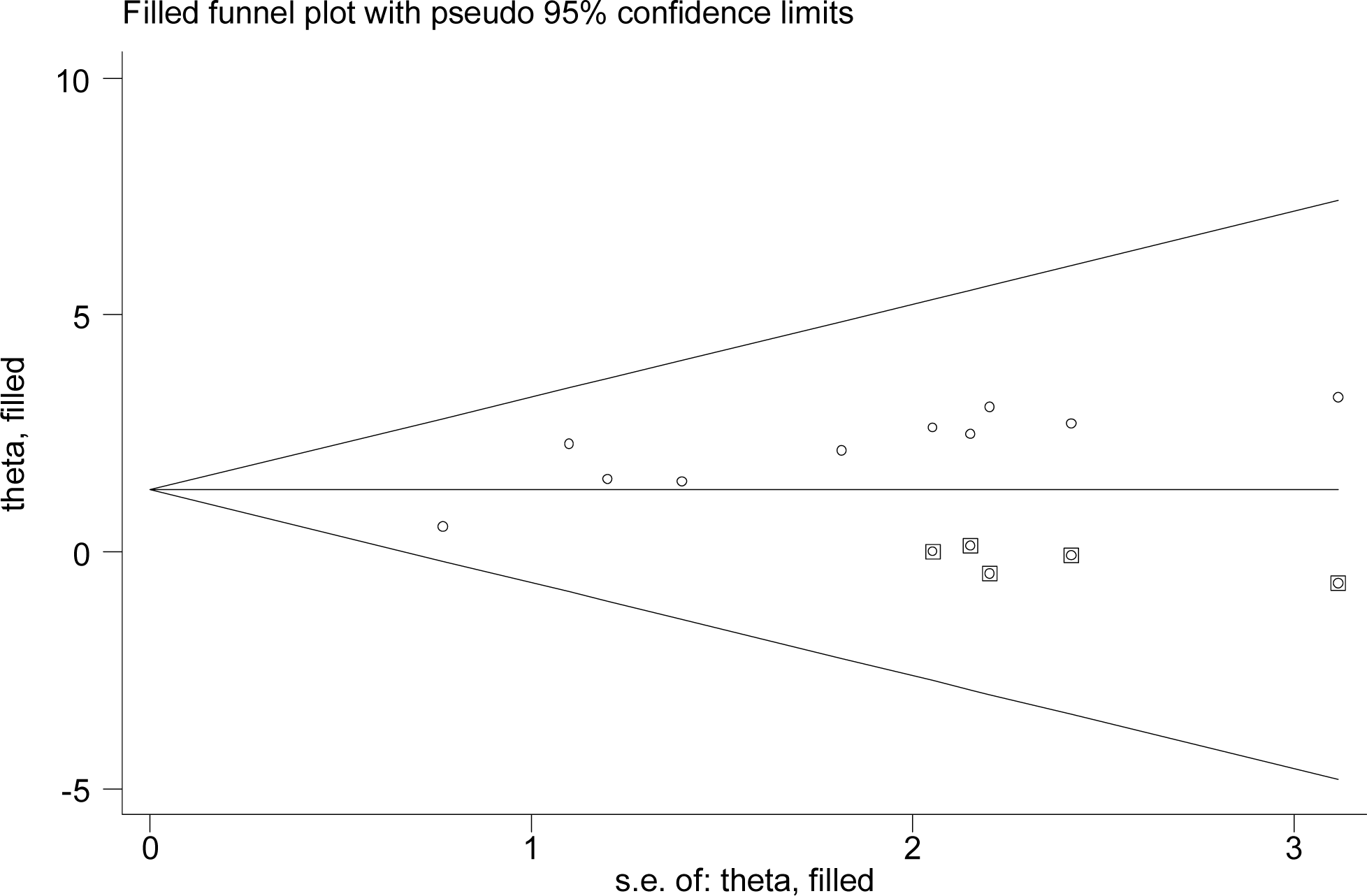
Result of trim and fill analysis after adjusting for publication bias of the 10 studies.

### Association diabetic foot ulcers with diabetic duration and patient residence

Diabetic patients with duration of diabetes more than 10 years were 3.91 times more likely to develop diabetic foot ulcers as compared with patients with duration of diabetes less than 10 years (OR, 3.91, 95% CI 2.03, 7.52, I^2^=63.4%) (Figure 5). Likewise, those diabetic patients who lived in the rural area were 3.40 times more likely to develop diabetic foot ulcers than those who lived in the urban area [OR = 3.40; 95% CI: 2.09, 5.54, I^2^=0.0%] (Figure 6).

**Figure 5:**
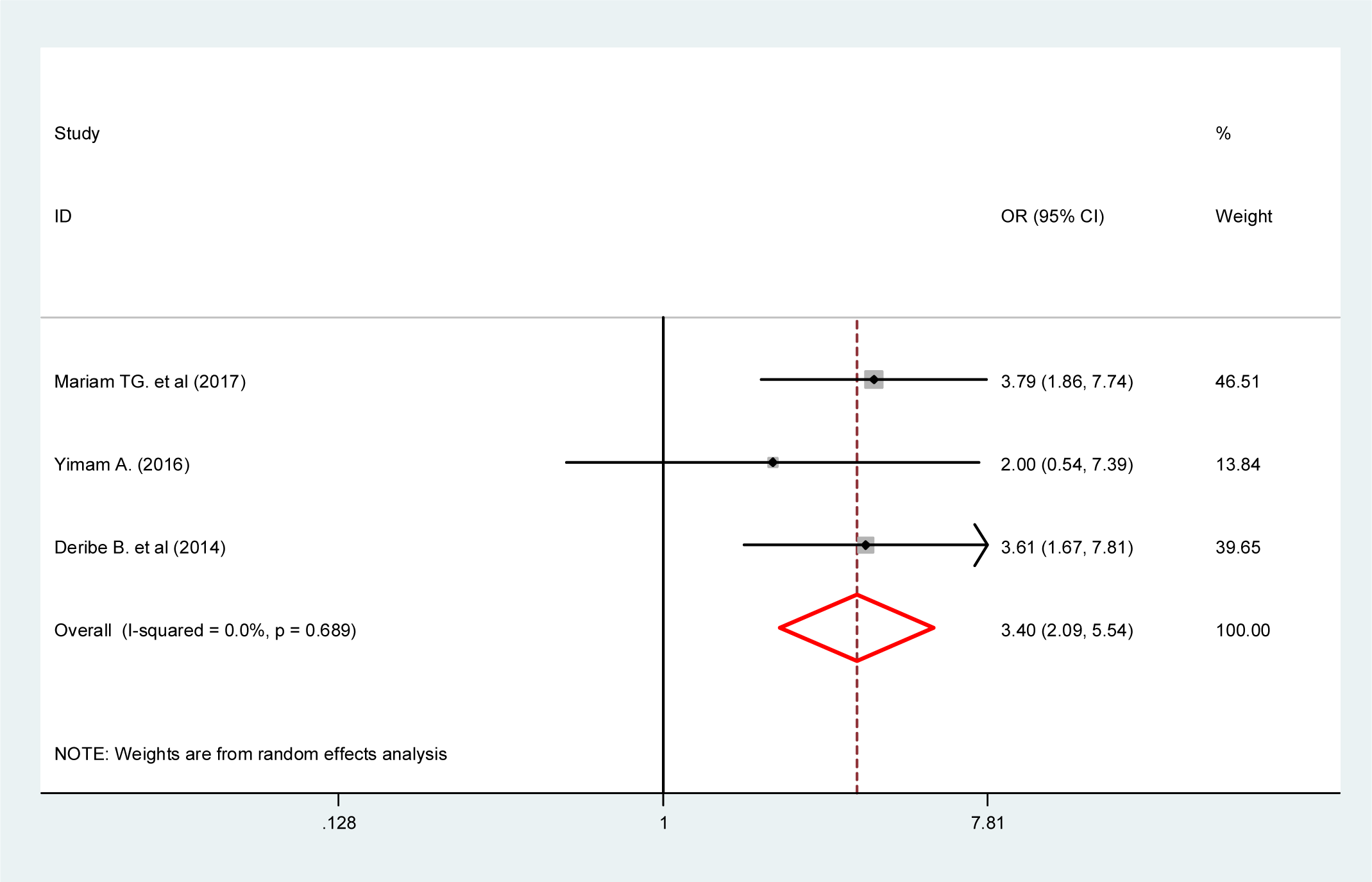
Forest plot showing the association of patient residence and diabetic foot ulcers.

**Figure 6:**
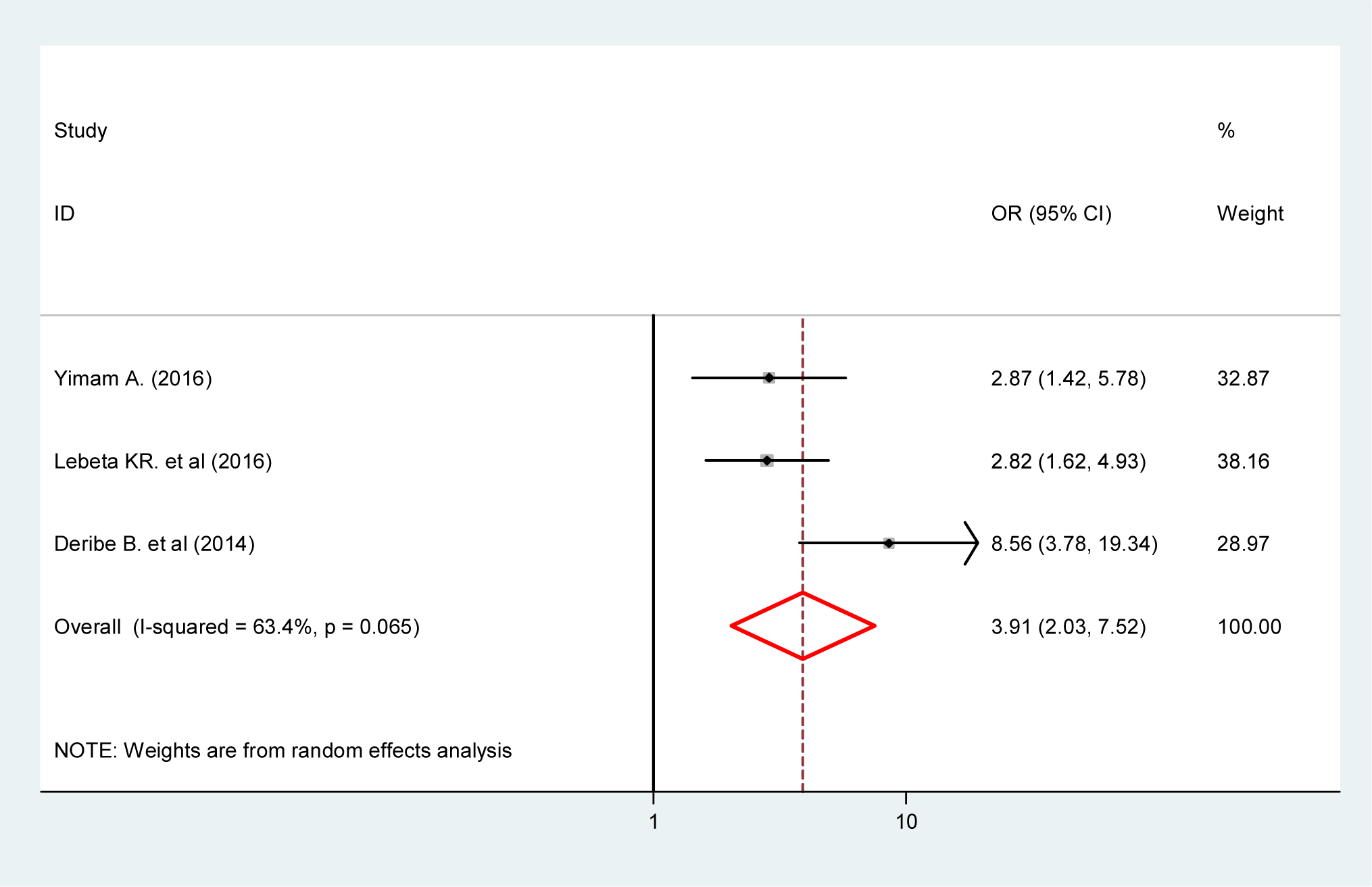
Forest plot showing the association of diabetic duration and diabetic foot ulcers.

## Discussion

The global prevalence of diabetic complications is rising rapidly[40]. Diabetic foot ulcer (DFU) is one of the major causes of morbidity, and it is responsible for 50% of diabetes-related hospital admissions in hospitals[41]. Moreover, a systematic literature review revealed that diabetes and related complications including foot ulceration are a common problem in Africa[42]. Early diagnosis and management of diabetic foot are critical to prevent its serious consequences such as infection, gangrene, and death[41].

In this study, the pooled national prevalence of diabetic foot among diabetic patients in Ethiopia was 11.27%. This finding was comparable with reports from other similar studies in Asia[43, 44] and Africa [45-47]. This might be due to similarities in the socio-demographic characteristics of the study participants. On the other hand, our finding was higher than the global (i.e. 6.3%) and national or continental (7.2% in Africa) prevalence of diabetic foot ulcers[19]. Likewise, our finding is higher than other similar studies[48, 49]. One possible explanation for the high prevalence of diabetic foot in Ethiopian diabetic patients might be due to variations in risk factors such as poor diabetic-related knowledge and poor foot self-care practice. In spite of that, the current study finding was lower than the study finding in Nigeria and Netherlands where the prevalence of diabetic foot ulcers was 41.5% and 20.4% respectively[50, 51]. It could be due to a difference in sample size and study setting.

Even though several factors have been reported as a risk factor of DFU in different literature[15, 44, 52-54], we have identified only two similar factors (duration of diabetes and patient residence) across the included studies for analysis. In agreement with studies conducted in Egypt, Iran and India[43, 54, 55], our study revealed that the duration of diabetes and patients’ residence had a significant association with a diabetic foot ulcer. Diabetic patients with long duration are exposed to multiple complications like neuropathy, which may increase the risk of foot ulceration due to trauma and high focal foot pressures[56]. Regarding residence, the possible reason might be since individuals in rural areas in Ethiopia are farmers and are often barefoot, which can result in damage to the foot and it leads to ulcers. Besides, most of the people in rural areas of the country are poor with low educational level so that they have no adequate knowledge and practice of foot care. However, the controversial result was reported from Sudan where diabetic patients from the urban area have been shown significantly associated with the development of DFU[57].

The findings of this meta-analysis have implications for clinical practice. Determining the prevalence of DFU provides up-to-date evidence to develop prevention, public health planning and management strategy for diabetic foot ulcers. It also used to improve patients’ quality of life and reduce the economic burden. Moreover, identifying determinant factors help clinicians to consider during their routine clinical practice. However, some limitations should be considered in future researches. It was difficult to analyze some important major factors since they were not examined consistently across the studies. Moreover, we identified significant heterogeneity across the included studies and the presence of publication bias so that the interpretation of the results has to be taken cautiously.

## Conclusion

Available data suggest that the prevalence of active DFU in people with diabetes in Ethiopia was high, but it is all derived from selected populations. Duration diabetes and Patients’ residence were significant factors for foot ulceration. Diabetes care in Ethiopia needs to develop a high standard of treatment guideline and preventive strategies to reduce the burden of DFU and prevent its risk factors. Health education should be given to patients with a longer duration of diabetes and those living in rural areas to reduce the progression of the disease and improve health outcomes. Moreover, there is a clear need for a large-scale prospective study to determine the extent of this important health care problem.

## Data Availability

The data analyzed during the current systematic review and meta-analysis is available from the corresponding author on reasonable request.

## Abbreviations

CI: Confidence Interval
FMoH: Federal Ministry of Health
OR: Odds Ratio
PRISMA: Preferred Reporting Items for Systematic Reviews and Meta-Analyses
SNNP: Southern Nations, Nationalities, and Peoples
DFUs: Diabetic Foot Ulcers

## Declarations

### Consent for publication

Not applicable.

### Ethics approval and consent to participate

Not applicable.

### Authors’ contributions

HM developed the protocol and involved in the design, selection of study, data extraction, and statistical analysis and developing the initial drafts of the manuscript. GD, FW, HZ, BT, and HB involved in data extraction, quality assessment, statistical analysis and revising subsequent drafts. HM, FW and TDH prepared the final draft of the manuscript. All authors read and approved the final draft of the manuscript.

## Acknowledgment

we would like to thank all authors of studies included in this systematic review and meta-analysis.

## References

1. Lakshmi N, Patel N, Parmar P, Garasiya K, Chaudhary M: Study the foot care practice among diabetic patients in Ahmedabad city, Gujarat. International Journal of Medical Science and Public Health 2018, 7(5): 333–338.

2. Teshome HM, Ayalew GD, Shiferaw FW, Leshargie CT, Boneya DJ: The Prevalence of Depression among Diabetic Patients in Ethiopia: A Systematic Review and Meta-Analysis, 2018. Depression Research and Treatment 2018, 2018.

3. Wild S, Roglic G, Green A, Sicree R, King H: Global prevalence of diabetes: estimates for the year 2000 and projections for 2030. Diabetes care 2004, 27(5): 1047–1053.

4. Hall V, Thomsen RW, Henriksen O, Lohse N: Diabetes in Sub Saharan Africa 1999-2011: epidemiology and public health implications. A systematic review. BMC public health 2011, 11(1): 564.

5. Alexiadou K, Doupis J: Management of diabetic foot ulcers. Diabetes Therapy 2012, 3(1): 4.

6. Pendsey SP: Understanding diabetic foot. International journal of diabetes in developing countries 2010, 30(2): 75.

7. Boulton AJ, Vileikyte L, Ragnarson-Tennvall G, Apelqvist J: The global burden of diabetic foot disease. The Lancet 2005, 366(9498): 1719–1724.

8. Singh N, Armstrong DG, Lipsky BA: Preventing foot ulcers in patients with diabetes. Jama 2005, 293(2): 217–228.

9. Agwu E, Dafiewhare EO, Ekanem PE: Possible diabetic-foot complications in Sub-Saharan Africa. In: Global Perspective on Diabetic Foot Ulcerations. edn.: InTech; 2011.

10. Levitt N: Diabetes in Africa: epidemiology, management and healthcare challenges. Heart 2008.

11. Jeffcoate WJ, Harding KG: Diabetic foot ulcers. The lancet 2003, 361(9368): 1545–1551.

12. Nabuurs-Franssen M, Huijberts M, Kruseman AN, Willems J, Schaper N: Health-related quality of life of diabetic foot ulcer patients and their caregivers. Diabetologia 2005, 48(9): 1906–1910.

13. Ribu L, Hanestad BR, Moum T, Birkeland K, Rustoen T: A comparison of the health-related quality of life in patients with diabetic foot ulcers, with a diabetes group and a nondiabetes group from the general population. Quality of Life Research 2007, 16(2): 179–189.

14. Kibachio J, Omolo J, Muriuki Z, Juma R, Karugu L, Ng’ang’a Z: Risk factors for diabetic foot ulcers in type 2 diabetes: A case control study, Nyeri, Kenya. African Journal of Diabetes Medicine 2013, 21(1).

15. Desalu O, Salawu F, Jimoh A, Adekoya A, Busari O, Olokoba A: Diabetic foot care: self reported knowledge and practice among patients attending three tertiary hospital in Nigeria. Ghana medical journal 2011, 45(2).

16. Lavery LA, Armstrong DG, Wunderlich RP, Mohler MJ, Wendel CS, Lipsky BA: Risk factors for foot infections in individuals with diabetes. Diabetes care 2006, 29(6): 1288–1293.

17. Lipsky BA, Berendt AR, Deery HG, Embil JM, Joseph WS, Karchmer AW, LeFrock JL, Lew DP, Mader JT, Norden C: Diagnosis and treatment of diabetic foot infections. Clinical Infectious Diseases 2004:885–910.

18. Wikblad K, Smide B, Bergström A, Kessi J, Mugusi F: Outcome of clinical foot examination in relation to self-perceived health and glycaemic control in a group of urban Tanzanian diabetic patients. Diabetes research and clinical practice 1997, 37(3): 185–192.

19. Zhang P, Lu J, Jing Y, Tang S, Zhu D, Bi Y: Global epidemiology of diabetic foot ulceration: a systematic review and meta-analysis. Annals of medicine 2017, 49(2): 106–116.

20. Wu SC, Driver VR, Wrobel JS, Armstrong DG: Foot ulcers in the diabetic patient, prevention and treatment. Vascular health and risk management 2007, 3(1): 65.

21. Liberati A, Altman DG, Tetzlaff J, Mulrow C, Gøtzsche PC, Ioannidis JP, Clarke M, Devereaux PJ, Kleijnen J, Moher D: The PRISMA statement for reporting systematic reviews and meta-analyses of studies that evaluate health care interventions: explanation and elaboration. PLoS medicine 2009, 6(7): e1000100.

22. Modesti PA, Reboldi G, Cappuccio FP, Agyemang C, Remuzzi G, Rapi S, Perruolo E, Parati G: Panethnic differences in blood pressure in Europe: a systematic review and meta-analysis. PloS one 2016, 11(1): e0147601.

23. McFarland LV: Meta-analysis of probiotics for the prevention of antibiotic associated diarrhea and the treatment of Clostridium difficile disease. The American journal of gastroenterology 2006, 101(4): 812.

24. Higgins JP, Thompson SG, Deeks JJ, Altman DG: Measuring inconsistency in meta-analyses. BMJ: British Medical Journal 2003, 327(7414): 557.

25. Islam MM, Iqbal U, Walther B, Atique S, Dubey NK, Nguyen P-A, Poly TN, Masud JHB, Li Y-CJ, Shabbir S-A: Benzodiazepine use and risk of dementia in the elderly population: a systematic review and meta-analysis. Neuroepidemiology 2016, 47(3-4):181–191.

26. Ried K: Interpreting and understanding meta-analysis graphs: a practical guide. 2006.

27. Tang J-L, Liu JL: Misleading funnel plot for detection of bias in meta-analysis. Journal of clinical epidemiology 2000, 53(5): 477–484.

28. Egger M, Smith GD, Phillips AN: Meta-analysis: principles and procedures. Bmj 1997, 315(7121): 1533–1537.

29. Adem A, Demis T, Feleke Y: Trend of diabetic admissions in Tikur Anbessa and St. Paul’s University Teaching Hospitals from January 2005-December 2009, Addis Ababa, Ethiopia. Ethiopian medical journal 2011, 49(3): 231–238.

30. Ejigu A: Brief communication: Patterns of chronic complications of diabetic patients in Menelik II Hospital, Ethiopia. Ethiopian Journal of health development 2000, 14(1): 113–116.

31. Yimam A: Prevalence od Diabetic foot ulcer and associated factors among diabetic patient attending Tikur Anbesa Specialized Hospital Diabetic clinic, Addis Ababa, Ethiopia, 2017. 2017.

32. Abejew AA, Belay AZ, Kerie MW: Diabetic complications among adult diabetic patients of a tertiary hospital in Northeast Ethiopia. Advances in Public Health 2015, 2015.

33. Kidist Reba Lebeta, Zeleke Argaw, Birhane BW: Prevalence of Diabetic Complications and Its Associated Factors Among Diabetes Mellitus Patients Attending Diabetes Mellitus Clinics; Institution Based Cross Sectional Study. American Journal of Health Research 2017, 5(2): 38–43.

34. Mariam TG, Alemayehu A, Tesfaye E, Mequannt W, Temesgen K, Yetwale F, Limenih MA: Prevalence of Diabetic Foot Ulcer and Associated Factors among Adult Diabetic Patients Who Attend the Diabetic Follow-Up Clinic at the University of Gondar Referral Hospital, North West Ethiopia, 2016: Institutional-Based Cross-Sectional Study. Journal of diabetes research 2017, 2017.

35. Tilahun A, Waktola C, Tewodros G, Sadik G, Amare D: Major Microvascular Complications and Associated Risk Factors among Diabetic Outpatients in Southwest Ethiopia. Endocrinol Metab Syndr 2017, 6(272): 2161-1017.10002.

36. Worku D, Hamza L, Woldemichael K: Patterns of diabetic complications at jimma university specialized hospital, southwest ethiopia. Ethiopian journal of health sciences 2010, 20(1).

37. Deribe B, Woldemichael K, Nemera G: Prevalence and factors influencing diabetic foot ulcer among diabetic patients attending Arbaminch Hospital, South Ethiopia. J Diabetes Metab 2014, 5(1): 1–7.

38. Gebrekirstos K, Gebrekiros S, Fantahun A: Prevalence and Factors Associated With Diabetic Foot Ulcer among Adult Patients in Ayder Referral Hospital Diabetic Clinic Mekelle, North Ethiopia, 2013. J Diabetes Metab 2015, 6(579): 2.

39. Duval S, Tweedie R: Trim and fill: a simple funnel-plot–based method of testing and adjusting for publication bias in meta-analysis. Biometrics 2000, 56(2): 455–463.

40. Susan van D, Beulens JW, Yvonne T. van der S, Grobbee DE, Nealb B: The global burden of diabetes and its complications: an emerging pandemic. European Journal of Cardiovascular Prevention & Rehabilitation 2010, 17(1_suppl):s3–s8.

41. Kasiya MM, Mang’anda GD, Heyes S, Kachapila R, Kaduya L, Chilamba J, Goodson P, Chalulu K, Allain TJ: The challenge of diabetic foot care: Review of the literature and experience at Queen Elizabeth Central Hospital in Blantyre, Malawi. Malawi Medical Journal 2017, 29(2): 218–223.

42. Bos M, Agyemang C: Prevalence and complications of diabetes mellitus in Northern Africa, a systematic review. BMC public health 2013, 13(1): 387.

43. Shahi SK, Kumar A, Kumar S, Singh SK, Gupta SK, Singh T: Prevalence of diabetic foot ulcer and associated risk factors in diabetic patients from North India. The Journal of Diabetic Foot Complications 2012, 4(3): 83–91.

44. Yusuf S, Okuwa M, Irwan M, Rassa S, Laitung B, Thalib A, Kasim S, Sanada H, Nakatani T, Sugama J: Prevalence and risk factor of diabetic foot ulcers in a regional hospital, eastern Indonesia. Open Journal of Nursing 2016, 6(01): 1.

45. Ndip EA, Tchakonte B, Mbanya J-C: A study of the prevalence and risk factors of foot problems in a population of diabetic patients in cameroon. The international journal of lower extremity wounds 2006, 5(2): 83–88.

46. Neuhann H, Warter-Neuhann C, Lyaruu I, Msuya L: Diabetes care in Kilimanjaro region: clinical presentation and problems of patients of the diabetes clinic at the regional referral hospital— an inventory before structured intervention. Diabetic medicine 2002, 19(6): 509–513.

47. Ogbera A, Fasanmade O, Ohwovoriole A, Adediran O: An assessment of the disease burden of foot ulcers in patients with diabetes mellitus attending a teaching hospital in Lagos, Nigeria. The international journal of lower extremity wounds 2006, 5(4): 244–249.

48. Nyamu P, Otieno C, Amayo E, McLigeyo S: Risk factors and prevalence of diabetic foot ulcers at Kenyatta National Hospital, Nairobi. East African Medical Journal 2003, 80(1): 36–43.

49. Walters D, Catling W, Mullee M, Hill R: The distribution and severity of diabetic foot disease: a community study with comparison to a non-diabetic group. Diabetic Medicine 1992, 9(4): 354–358.

50. Bouter K, Storm A, Uitslager R, Erkelens D, Diepersloot R: The diabetic foot in Dutch hospitals: epidemiological features and clinical outcome. The European journal of medicine 1993, 2(4): 215–218.

51. Ogbera A, Adedokun A, Fasanmade O, Ohwovoriole A, Ajani M: The foot at risk in Nigerians with diabetes mellitus-the Nigerian scenario. International journal of Endocrinology and Metabolism 2005, 2005(4, Autumn):165-173.

52. Adler AI, Boyko EJ, Ahroni JH, Stensel V, Forsberg RC, Smith DG: Risk factors for diabetic peripheral sensory neuropathy: results of the Seattle Prospective Diabetic Foot Study. Diabetes care 1997, 20(7): 1162–1167.

53. Ahmad W, Khan IA, Ghaffar S, Al-Swailmi FK, Khan I: Risk factors for diabetic foot ulcer. Journal of Ayub Medical College Abbottabad 2013, 25(1-2):16–18.

54. Yazdanpanah L, Shahbazian H, Nazari I, Arti HR, Ahmadi F, Mohammadianinejad SE, Cheraghian B, Latifi SM: Prevalence and related risk factors of diabetic foot ulcer in Ahvaz, south west of Iran. Diabetes & Metabolic Syndrome: Clinical Research & Reviews 2018.

55. El Din SA, Mekkawy MM, Besely WN, Azer SZ: Prevalence of Risk Factors for Egyptian Diabetic Foot Ulceration. Journal of Nursing a nd Health Science 2016; 5 (2): 45, 57.

56. Frykberg RG, Zgonis T, Armstrong DG, Driver VR, Giurini JM, Kravitz SR, Landsman AS, Lavery LA, Moore JC, Schuberth JM: Diabetic foot disorders: a clinical practice guideline (2006 revision). The journal of foot and ankle surgery 2006, 45(5): S1–S66.

57. Awadalla H, Noor SK, Elmadhoun WM, Almobarak AO, Elmak NE, Abdelaziz SI, Sulaiman AA, Ahmed MH: Diabetes complications in Sudanese individuals with type 2 diabetes: Overlooked problems in sub-Saharan Africa? Diabetes & Metabolic Syndrome: Clinical Research & Reviews 2017, 11:S1047–S1051.

